# Regional Genetic Architecture of the Corpus Callosum and its Overlap with Psychiatric and Substance Use Phenotypes

**DOI:** 10.1101/2023.12.03.23299351

**Authors:** Megan L. Campbell, Alexey Shadrin, Dennis van der Meer, Markos Tesfaye, Olav B. Smeland, Kevin S. O’Connell, Nadine Parker, Oleksandr Frei, Ole Andreassen, Dan J. Stein, Shareefa Dalvie, Jaroslav Rokicki

## Abstract

**Background:** The corpus callosum (CC) is a brain structure with a high heritability and a potential role in psychiatric and substance use traits. However, the genetic relationship of the CC, and its subregions, with psychiatric phenotypes remains largely unclear. We aimed to investigate the genetic architecture of the CC and its subregions and its overlap with psychiatric phenotypes using causal mixture model analysis.

**Methods:** We employed summary statistics from genome-wide association studies (GWAS) of the CC (n=35,592), psychiatric- (attention deficit hyperactivity disorder, ADHD; autism spectrum disorder, ASD; bipolar disorder, BD; major depressive disorder, MDD; schizophrenia, SCZ) and substance use- phenotypes (drinks per week, smoking cessation, smoking initialization, number of cigarettes per day and cannabis use) phenotypes, and performed bivariate casual mixture model analyses with MiXeR to investigate the genetic overlap between these phenotypes. Conjunctional false discovery rate (conjFDR) was used to identify genetic loci that exhibit joint associations with the CC and psychiatric or substance use phenotypes. Open Targets was used to annotate significant, independent variants and MAGMA was used to conduct gene-set enrichment analyses.

**Results:** The polygenicity of the CC subregions ranges from 1.4k variants in the posterior subregion to 1.8k variants in the mid posterior subregion, with approximately 1.8k variants influencing total CC volume. The extent of genetic overlap between CC and psychiatric and substance use phenotypes varied across CC subregion; loci contributing to anterior regions showing almost complete overlap with those involved in BD and SCZ (96% and 98%, respectively), while a distinct pattern emerged for MDD indicating fewer variants overlapping with central and mid anterior subregions (47% and 38%, respectively), loci contributing to the central region exhibited substantial overlap with drinks per week (87%). Loci contributing to the CC subregions exhibited lower overlap with ASD and ADHD (26% to 48%). The conjunctional analysis of CC subregions and psychiatric phenotypes revealed a total of 138 significant independent variants, with gene-set enrichment analyses implicating antigen processing, regulation of the extracellular matrix and hyaluronan metabolism.

**Conclusion:** We observed significant genetic overlap between the CC and SCZ, BD, and drinks per week, identifying loci linked to brain-related traits with specific overlap patterns found across conditions. Overlapping genetic architectures suggest shared biological pathways, and the potential value of targeting such pathways for the purposes of treatment.

## Introduction

The corpus callosum (CC) is the largest white matter tract in the brain and serves as the major interhemispheric commissure, integrating information from the two cerebral hemispheres to facilitate language, affective and sensorimotor function (Wang et al., 2020). The CC is commonly segmented into five subregions: anterior, mid-anterior, central, mid-posterior and posterior, with each subregion having distinct axonal projections from cortical regions (Fischl et al., 2002; Hofer and Frahm, 2006; Witelson, 1989). Previous work examining the heritability of the volume of CC and its subregions has revealed a polygenic architecture, and suggested genetic overlap with neuropsychiatric disorders. However, the extent to which the genetic architecture of the volume of CC overlaps with common neuropsychiatric- and substance use phenotypes is largely unknown (Campbell et al., 2023; Gadewar et al., 2023).

Neuroimaging studies have provided insight into the structural and functional differences in the CC among individuals with various neuropsychiatric and substance use disorders. Studies in bipolar disorder (BD) have identified altered white matter integrity in the CC, potentially affecting interhemispheric communication (Wise et al., 2016). Furthermore, individuals with schizophrenia (SCZ) show structural abnormalities such as reduced thickness and altered shape in the CC, indicating disrupted connectivity between brain regions (Frumin et al., 2002; Lu et al., 2011). In BD and SCZ, disrupted interhemispheric communication may contribute to the cognitive and emotional dysregulation observed in these disorders (Frumin et al., 2002; Wise et al., 2016). In individuals with attention deficit hyperactivity disorder (ADHD), there is evidence of decreased volume and alterations in the microstructure of the CC, indicating disruptions in interhemispheric connectivity (Acer et al., 2017). Similarly, individuals with autism spectrum disorder (ASD) exhibit differences in CC size, shape, and connectivity, suggesting atypical development of white matter microstructure. Substance use-related traits have also been associated with alterations in CC volume, but findings regarding the effects of alcohol consumption, cigarette smoking, and cannabis use on CC volume are inconsistent (Hampton et al., 2019; Moeller et al., 2004).

Despite the known genetic contributions to both the CC and to psychiatric and substance use phenotypes, the extent of genetic overlap of the CC and its subregions with these phenotypes remains unclear. We previously found significant genetic correlations between total CC volume, BD and drinks per week, suggestive of genetic overlap between these traits (Campbell et al., 2023). However, genome-wide genetic correlations do not fully characterize genetic overlap since they cannot detect an overlap when shared trait-influencing variants have a balanced mixture of concordant and opposing directions of effect, thus cancelling out genome-wide correlation. In this study, our aims are twofold: first, we aim to provide a comprehensive description of the genetic architecture of the CC, by investigating the polygenicity determining the number of trait influencing variants. Second, we seek to further delineate the relationship between the CC and relevant phenotypes, utilizing the causal mixture model framework (MiXeR) to quantify genetic overlap independent of genetic correlations (Frei et al., 2019). Additionally, we employ conjunctional false discovery rate (conjFDR) to identify loci jointly associated with both phenotypes (Andreassen et al., 2013). This approach allows us to gain a more nuanced understanding of the shared genetic components between the CC and psychiatric phenotypes, while addressing the limitations of conventional genetic correlation analyses.

## Methods

### Genome-wide Association Studies

We compared genome-wide association studies (GWAS) of five subregions of the CC and total CC volume (conducted using data from the UK Biobank (accession code 27412; Campbell et al., 2023) with GWAS of the following psychiatric phenotypes: ADHD (Demontis et al., 2019), ASD (PGC ASD, 2017), BD (Mullins et al., 2021), MDD (Wray et al., 2018), SCZ (Trubetskoy et al., 2022) and substance use traits: drinks per week (N=941,280), number of cigarettes per day (N=337,334), smoking cessation (N=547,219) and smoking initiation (N=1,232,091) (Liu et al., 2019) and cannabis use (Pasman et al., 2018). In all GWAS datasets except that of the CC, we excluded samples from the UK Biobank from phenotypes to avoid potential sample overlap as required by conjFDR. This was not possible for the GWAS of cannabis use as it consists predominantly of individuals from the UK Biobank. Since MiXeR accounts for sample overlap, we used the whole samples without the removal of overlapping samples. A full table of these GWAS can be found in the Supplementary Material (SM), Table 1.

### Analysis of genetic architecture using causal mixture modelling

Univariate MiXeR analyses were conducted to estimate the number of single nucleotide polymorphisms (SNPs) influencing each trait, accounting for linkage disequilibrium (LD). To investigate the genetic overlap between CC volume and the aforementioned phenotypes, causal mixture models were applied using MiXeR. Bivariate MiXeR analyses were performed to estimate the number of SNPs shared between each pair of phenotypes and the number of trait-specific SNPs associated with either of these phenotypes. For each univariate and bivariate MiXeR analysis 20 independent runs were performed using randomly selected SNPs. We report the average number of trait-influencing variants and corresponding standard deviations across these 20 runs. From the number of trait influencing variants, we calculated the percentage of SNPs (with respect to the number of shared variants) with the same direction of effect (concordant) and those with opposite direction of effect (discordant). The difference between the Akaike Information Criterion (AIC) of the comparative model and that of the fitted model was calculated to serve as a measure of the relative model fit. A positive AIC difference indicates that the fitted model is suitable, indicating better model discrimination. (Hindley et al., 2022). Additionally, model fit was evaluated based on predicted versus observed conditional quantile-quantile plots (QQ-plots) and log-likelihood plots that visualize the parameter estimation procedure (SM Table 2).

### Identification of shared genetic loci with conjunctional false discovery rate

To identify loci associated with the CC and those associated with conditional phenotypes, we employed the pleiotropy-informed conjFDR method (accessed at https://github.com/precimed/pleiofdr; Andreassen et al., 2013). The conjFDR combines GWAS summary statistics of two phenotypes to identify genetic loci that exhibit joint associations with both phenotypes. The conjFDR statistic, calculated from two conditional FDR statistics obtained by swapping the roles of primary and conditional phenotypes, identifies SNPs associated with both the primary and the conditional phenotypes. It yields an estimate of the posterior probability that a SNP is null for either or both traits, provided that the p-values for both phenotypes combined are equal to or smaller than the p-values for each individual trait. A conjFDR threshold of 5% was applied to identify statistically significant loci associated with these phenotypes.

SNPs within the extended major histocompatibility complex (MHC, GRCh37: 6:26,000,000–33,000,000) region were excluded prior to fitting the conjFDR models to mitigate potential bias due to the intricate LD structure of the MHC. Random pruning of SNPs in 500 iterations was performed to avoid the inflation of the results due to LD-dependency. For every iteration a single SNP was randomly selected to represent each LD block (clump of variants with an r^2^ value greater than 0.1), and their respective p-values were used to compute the empirical cumulative distribution functions. The results were then averaged across all iterations.

### Functional annotation of shared loci

Open Targets Genetics was used to annotate all significant, independent SNPs (Mountjoy et al., 2021). Open Targets maps SNPs to genes by combining positional information regarding the distance between the variant and each genes canonical transcription start site, expression quantitative trait loci (QTL), protein QTL, splicing QTL, epigenomic data, and functional prediction (Mountjoy et al., 2021). Open Targets integrates this information and calculates variants Locus2Gene score, which enables the identification of the gene with the strongest association with the variant. All variants in the major histocompatibility complex (MHC) region (GRCh37: 6:26,000,000–33,000,000) were excluded prior to annotation. Following annotation, the Gene2Func function in Functional Mapping and Annotation of GWAS (FUMA, platform v1.3.7, accessed October 2023; (Watanabe et al., 2017)) was used to conduct gene-set enrichment analyses, utilising MAGMA v.1.08 (http://ctg.cncr.nl/software/magma; (de Leeuw et al., 2015). Genes were investigated for enrichment of biological processes, tissue and cell types.

## Results

### Analysis of genetic architecture using causal mixture modelling

In univariate MiXeR analysis, we estimated that CC volume has approximately 1.8×10^3^±88 trait influencing variants (here and below reported numbers of trait-influencing variants are truncated to variants with strongest effects which cumulatively account for 90% of phenotype’s heritability) (Figure 1). The numbers of trait influencing variants for the CC subregions were similar: anterior (1.7×10^3^±145), mid anterior (1.6×10^3^±90), central (1.5×10^3^±109), mid posterior (1.8×10^3^±124) and posterior (1.4×10^3^±70). Additionally, we found the CC to be less polygenic than substance use phenotypes: drinks per week 6.4×10^3^±508, smoking cessation 2.6×10^3^±465, smoking initiation 9.6×10^3^±316, cannabis consumption 8.1×10^3^±911 and cigarettes per day 0.9×10^3^±475 and psychiatric phenotypes: ADHD 3.9×10^3^±158, ASD 1.2×10^4^±1590, BD 8.6×10^3^±307, MDD 2.1×10^4^±2323, SCZ 9.6×10^3^±254.

**Figure 1:**
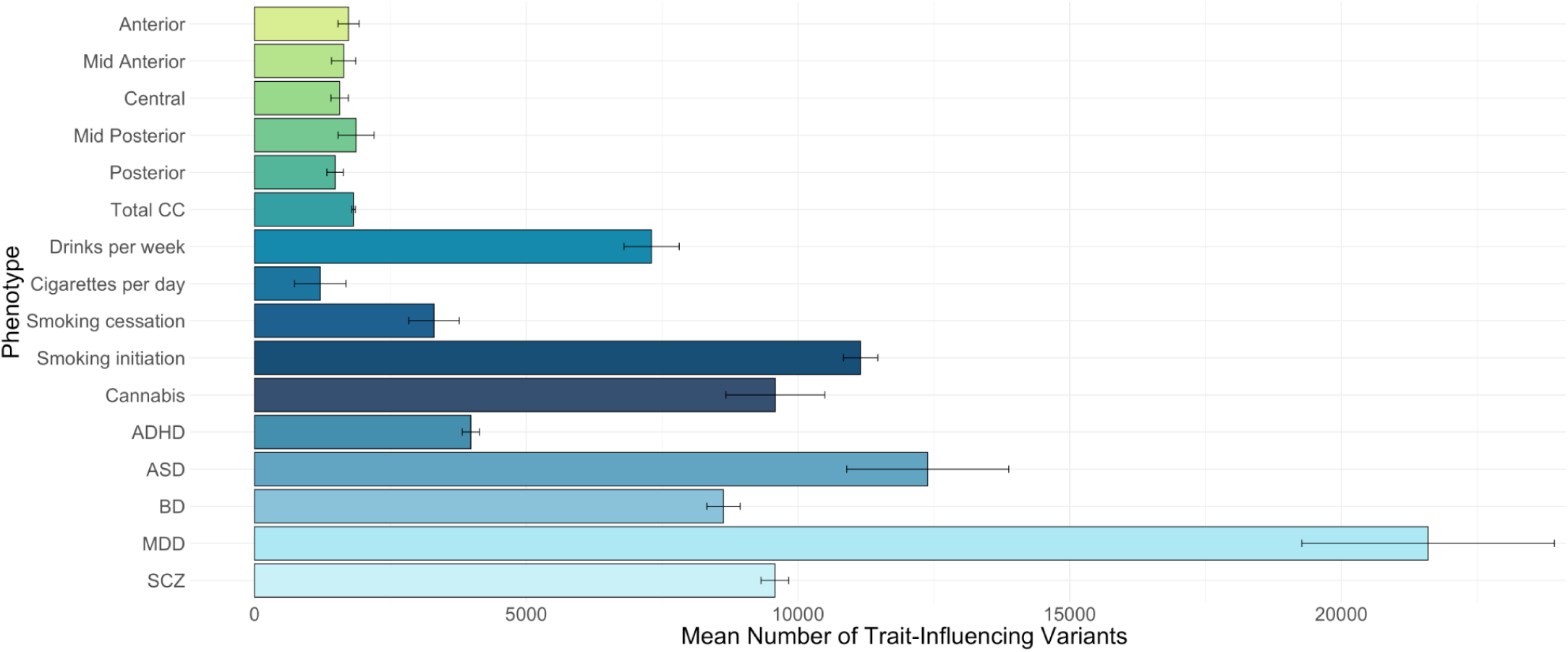
Polygenicity of the CC and psychiatric phenotypes illustrated by estimated causal variants. Polygenicity of volumes of the corpus callosum (CC) subregions (anterior, mid anterior, central, mid posterior, posterior), total CC volume, drinks per week, cigarettes per day, smoking cessation, smoking initiation, cannabis, attention deficit hyperactivity disorder (ADHD), autism spectrum disorder (ASD), bipolar disorder (BD), major depressive disorder (MDD), schizophrenia (SCZ).

The mid anterior and anterior subregions consistently showed a greater proportion of shared variants with psychiatric disorders, ranging from 91% to 97% for the anterior subregion (percentage calculated with respect to the CC) compared to other CC subregions (Figure 2). BD and SCZ exhibited similar patterns of overlap with the CC, with the highest proportion of overlap observed in the anterior subregion (91% and 97%, respectively) and the lowest in the mid posterior subregion for both disorders (20% and 15%, respectively). Ninety-five percent of the variants influencing the anterior subregion overlapped with MDD. However, MDD had a lower proportion of overlap with the central and mid anterior subregions (47% and 38%, respectively). For both ADHD and ASD, the proportion of overlapping variants for all subregions was less than 50%, except for ASD and the anterior subregion, which exhibited a 95% overlap in shared causal variants. When examining the directions of SNP effects in shared causal variants, only BD and ASD displayed more than 50% of SNPs with a consistent direction of effect. Specifically, 80% of the shared causal variants between the mid posterior subregion and BD had a consistent direction of effect, while 64% of the shared variants between the posterior subregion and ASD exhibited concordance.

**Figure 2:**
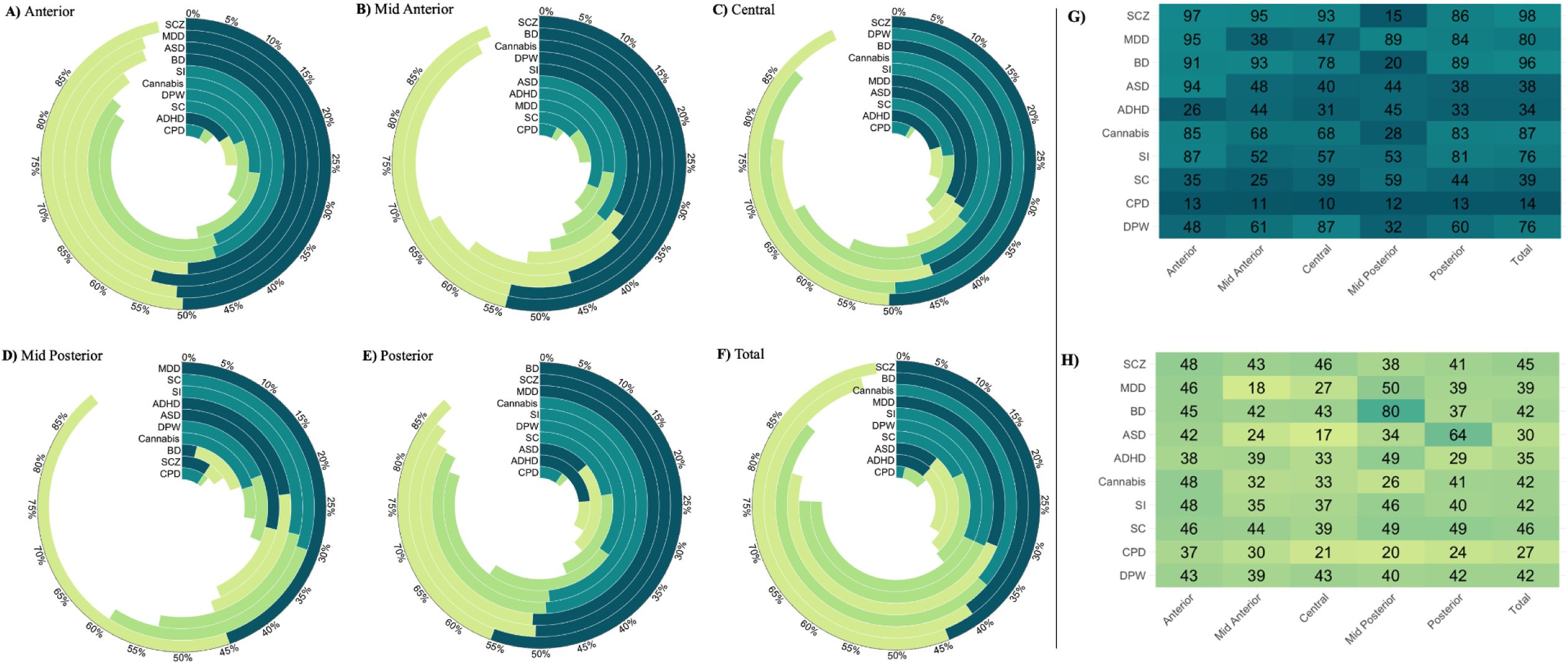
Shared trait influencing variants of the CC and psychiatric phenotypes. A-F) The percentage of concordant/ discordant SNPs from the number of shared trait influencing variants between variants (percentages are calculated with respect to total CC variants) G) Heatmap illustrating the estimated percentage of shared variants, calculated with respect to the CC H) Proportion of shared variants, showing the same direction of effect

In contrast to the psychiatric disorders, the substance use phenotypes exhibited a more variable genetic overlap with the CC and its subregions. Drinks per week had its highest overlap with the central subregion (87%), with lower overlap noted in the mid anterior and posterior subregions (61% and 60%, respectively) and the lowest proportion of overlap with the mid posterior subregion (32%). Cigarettes per day had the lowest proportion of shared variants, compared to the other psychiatric phenotypes, across all CC subregions (< 20. However, smoking cessation showed slightly higher overlap across subregions compared to cigarettes per day, with the highest overlap in the mid posterior subregion at 59%. Smoking initiation had a higher degree of overlap across all subregions, with the highest overlap in the anterior and posterior subregions (87% and 81%, respectively); the mid posterior, central, and mid anterior subregions had lower overlap, all below 60%. Among the substance use phenotypes, cannabis use had the highest overlap across the CC, with the highest overlap in the anterior subregion (85%) and the lowest in the mid posterior subregion (28%). In terms of SNP effects, the majority of shared causal variants were discordant (>50% discordant), across all comparisons. For cigarettes per day, across all CC subregions, over 70% of the shared variants had discordant directions of effect. Drinks per week, smoking cessation, and smoking initiation exhibited a similar pattern, with approximately 60% of variants indicating mixed effect directions across all CC subregions. For variants shared between cannabis use and the mid posterior, central and mid anterior subregions, ∼65% exhibited discordant directions of effect while slightly lower rates of discordance were noted for the anterior and posterior subregions (52% and 59%, respectively) (Figure 2A-F, H).

### Identification of shared genetic loci with conjunctional false discovery rate

We conducted conjFDR analysis using volumes of the CC and relevant psychiatric phenotypes to identify genetic loci shared between these phenotypes. The conjunctional analysis of CC subregions and psychiatric phenotypes revealed 110 significant loci (37 ADHD, 7 ASD, 44 BD, 6 MDD, 165 SCZ; conjFDR<0.05; SM Table 4) (Figure 3).

**Figure 3:**
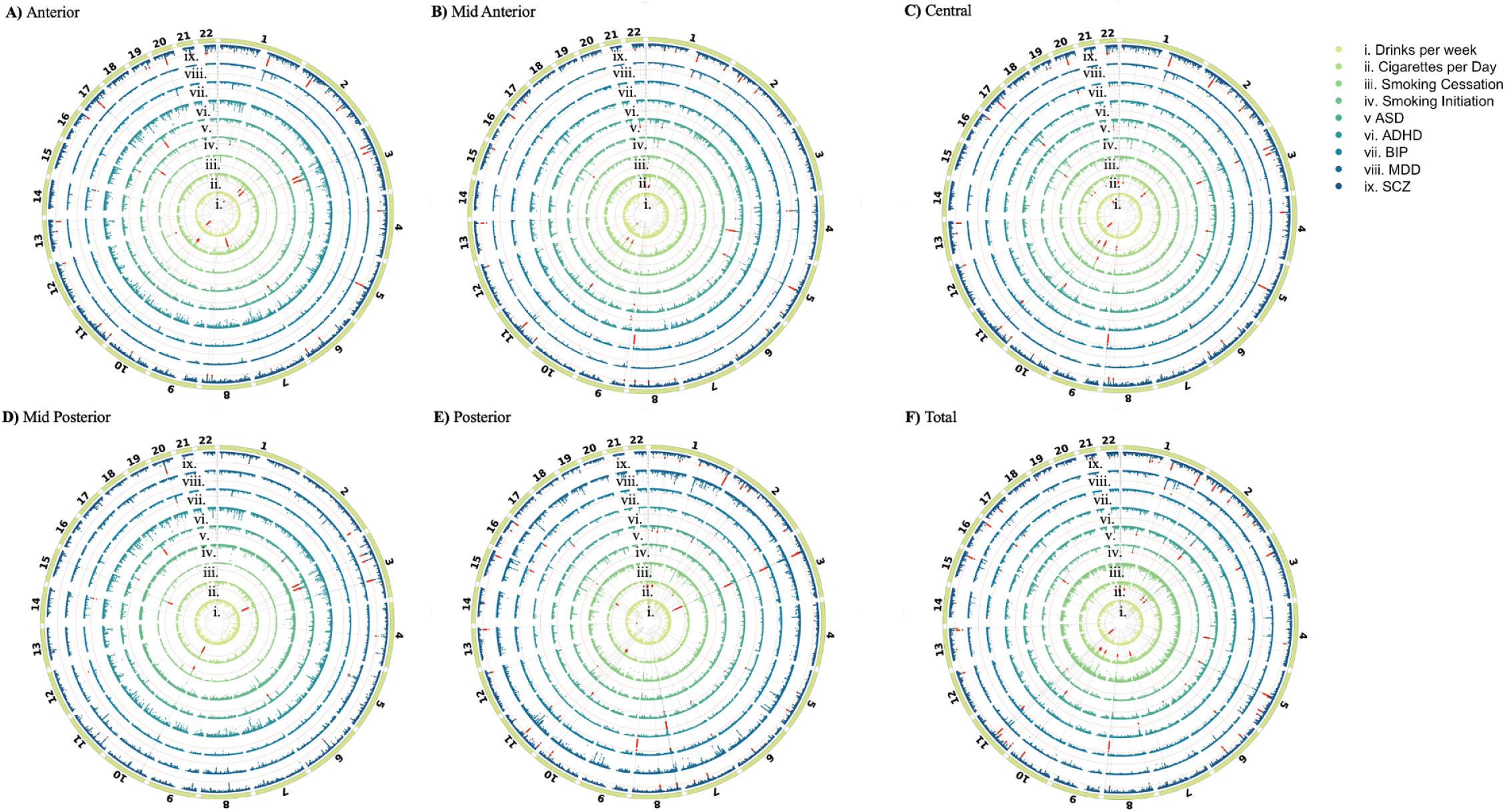
Manhattan plots of conjFDR results illustrated by subregion. A-F) ConjFDR results by subregion. Red dots indicated significant loci with FDR < 0.05. Significance level is indicated by dashed red line. Manhattans are plotted in the same order from inner circle to outer circle: Drinks per week, Cigarettes per day, Smoking cessation, Smoking initiation, ASD, Autism spectrum disorders; ADHD, Attention deficit hyperactivity disorder; BD, Bipolar disorder; MDD, Major depressive disorder; SCZ, Schizophrenia.

Twenty-eight unique loci were identified between the CC and substance use phenotypes (conjFDR<0.05) (4 drinks per week, 6 cigarettes per day, 20 smoking initiation, 2 smoking cessation; SM Table 5).

### Functional annotation of shared loci

Open Targets Genetics was used to annotate significant, independent SNPs (Mountjoy et al., 2021). Annotated genes were then used as the input for the Gene2Func function in FUMA to conduct gene-set enrichment analyses. All loci identified in the conjDFR analyses were intergenic or intronic SNPs. Only three comparisons had significant biological processes implicated from the gene-set analyses (SM Table 6): anterior subregion and ADHD (39 processes), central subregion and smoking initiation (5 processes), the posterior subregion and SCZ (1 process) and total CC volume and SCZ (4 processes). The top biological processes from ADHD and the anterior subregion implicated antigen processing (p=7×10^-7^; Bonferroni corrected). Negative regulation of the extracellular matrix was implicated by the central subregion and smoking initiation (p<0.05; Bonferroni corrected) and hyaluronan metabolism was implicated in both the posterior subregion and SCZ and total CC volume and SCZ.

## Discussion

We found the CC to be less polygenic than the psychiatric phenotypes we examined here, with approximately ∼1.8×10^3^ variants influencing total CC volume, and ∼1.4×10^3^ (posterior) to ∼1.8×10^3^ (mid-posterior) influencing CC subregions. The extent of genetic overlap between CC and psychiatric phenotypes varied across CC subregion; variants contributing to anterior regions showed almost complete overlap with those involved in BD and SCZ, while a distinct pattern emerged for MDD, indicating fewer variants overlapping with central subregions, variants contributing to the central region overlapped with alcohol consumption. Finally, we note similar patterns of genetic overlap between SCZ, BD and cannabis use with the CC.

Cannabis use and SCZ exhibited the similar patterns of overlap with the CC, emphasizing the complex interplay between these phenotypes. The observed genetic overlap here suggests shared risk variants, potentially influencing how individuals may be predisposed to both conditions (Vaissiere et al., 2020). Cannabis use has also been associated with changes in the CC microstructural organization, particularly in patients with first episode psychosis (Rigucci et al., 2016). Disruptions in this structure, influenced by genetics but exacerbated by cannabis use, may underlie some of the cognitive dysfunction inherent to SCZ.

The finding that BD and SCZ have similar genetic overlap with the anterior subregion, is consistent with a range of previous work. First, it is increasingly clear that there is extensive genetic overlap between BD and SCZ (Hindley et al., 2022; Smeland et al., 2020). Second, the anterior subregions of the CC play a crucial role in connecting the prefrontal cortices, areas integral to emotion regulation. Disruptions in the connectivity of this region could contribute to the observed emotional dysregulation inherent to psychotic spectrum disorders (Wise et al., 2016).

The mid-posterior subregion had the lowest number of overlapping trait-influencing variants with all traits. However, a notable exception emerges in the case of MDD, where there is almost complete genetic overlap with this region. Tracts originating from the mid-posterior subregion terminate in the occipital lobe, which has been consistently implicated in individuals with MDD (Kihira et al., 2020; Maciąg et al., 2010; Song et al., 2021). It is possible that changes in the connectivity or integrity of this subregion could contribute to the differences noted in the occipital lobe in individuals with MDD and could underlie some of the visual processing deficits and altered neurochemistry noted in the disorder.

It is noteworthy that there was limited genetic overlap contributing to CC with those contributing to ADHD and ASD. Although individuals with ASD have been shown to have reduced overall CC volume, the lack of evidence to suggest genetic overlap is in line with prior work (Campbell et al., 2023; Fame et al., 2011). The lower number of significant loci identified after conditioning on ADHD and ASD, may be due to the power of these GWAS. It is worth noting that MiXeR is robust to power fluctuations, suggesting that the observed smaller number of loci still indicates overlap. MiXeR reliably captures subthreshold signals, reinforcing the potential genetic connection between ADHD, ASD and the CC. It is worth to highlight this observation, especially as GWAS sample sizes constantly keep increasing, as this may enhance the overlap over time.

The central subregion presents a distinct pattern, with a consistently lower proportion of overlap with all substance use phenotypes, except for drinks per week, which has its highest proportion of overlap within this region. Of these shared variants, only 43% had a concordant direction of effect- indicating mixed genetic effects between the central subregion and alcohol consumption. These findings are consistent with work showing that increased alcohol consumption is associated with smaller central subregion volumes (González-Reimers et al., 2019). While the overall overlap of CC and substance use phenotypes is slightly lower than in psychotic disorders, there was substantial overlap with anterior and posterior subregions. This trend is most pronounced in the case of cannabis and smoking initiation traits.

Our gene-set enrichment analyses revealed a noteworthy association between the extracellular matrix (ECM) in both SCZ and smoking initiation and the CC. The ECM, a complex network of macromolecules, plays a pivotal role in maintaining the structural integrity of the CC. In the conjunctional analysis of SCZ and the CC, our findings implicated negative regulation of the ECM, aligning with previous studies that have reported alterations in ECM components in patients with SCZ (Pantazopoulos et al., 2020). These alterations can lead to neurophysiological dysregulation and progressive neuronal impairment. Our conjunctional analyses of the CC and smoking initiation, highlighted the significance of hyaluronan metabolism, a key component of the ECM. Hyaluronan contributes to the ECM’s viscoelastic properties, influencing cell behaviour and potentially modulates the neurobiological mechanisms underlying smoking initiation. These findings underscore the potential importance of the ECM in the pathophysiology of both SCZ and smoking initiation, and its role in the structural and functional dynamics of the CC. Further research into these associations could provide valuable insights into potential therapeutic targets.

Some limitations deserve emphasis. We used summary statistics from predominantly European GWAS, which limits the generalizability of these findings. Further work is needed with more diverse samples, and trans-ancestry tools. Additionally, since the conjunctional analyses require non-overlapping samples, we were not able to execute the analyses for all phenotypes (e.g. cannabis consumption). Additionally, some of our null findings may be due to low power in the original GWAS which may underestimate the shared genetic architecture between phenotypes.

In conclusion, we describe distinct patterns of genetic overlap, shedding light on the potential shared biological pathways and mechanisms underlying these associations. The genetic overlap identified between the CC, BD and SCZ aligns with previous research highlighting extensive genetic commonalities between these psychiatric disorders. The involvement of the anterior CC in emotion regulation and connectivity to prefrontal cortices further underscores the significance of these findings in understanding emotional dysregulation in psychotic spectrum disorders. Although replication is needed, overlapping genetic architectures suggest the involvement of shared biological pathways, and the potential value of targeting such pathways for the purposes of treatment.

## Data Availability

All data produced in the present study are available upon reasonable request to the authors, upon publication

## Notes

### Competing Interest Statement

The authors have declared no competing interest.

### Funding Statement

This study did not receive any funding

### Author Declarations

University of Cape Town Faculty of Health Sciences Human Research Ethics Committee (HREC)

